# The cost-effectiveness of thoracic epidural versus paravertebral blockade in reducing chronic post-thoracotomy pain – a trial-based economic evaluation

**DOI:** 10.1101/2025.08.07.25333206

**Authors:** Mishal Javed, Louise Jackson, Lee Middleton, Benjamin Shelley, Hannah Summers, Rebecca Boyles, Michael Gilbert, Andreas Goebel, Ira Goldsmith, Stephen Grant, Sajith Kumar, Nandor Marczin, Philip McCall, Rajnikant Mehta, Teresa Melody, Babu Naidu, Sridhar Rathinam, Lajos Szentgyorgyi, Sarah Tearne, Ben Watkins, Matthew Wilson, Andrew Worrall, Joyce Yeung, Fang Gao Smith

**Affiliations:** School of Health Sciences, University of Birmingham; Birmingham Clinical Trials Unit, University of Birmingham; University of Glasgow Anaesthesia, Perioperative Medicine and Critical Care Research Group and Department of Cardiothoracic Anaesthesia, Golden Jubilee National Hospital, Clydebank; Glenfield Hospital, University Hospitals of Leicester; Department of Anaesthetics, Morriston Hospital; Pain Research Institute, University of Liverpool, ILCAMS, Clinical Sciences Centre, and Liverpool and Walton Centre NHS Foundation Trust, Liverpool; Department of Cardiothoracic Surgery, Morriston Hospital, and Faculty of Medicine, Health and Life Science, Swansea University; Patient and Public Partner; University Hospitals Birmingham NHS Foundation Trust; Imperial College London, and Royal Brompton and Harefield NHS Foundation Trust; University of Glasgow Anaesthesia, Perioperative Medicine and Critical Care Research and Department of Cardiothoracic Anaesthesia, Golden Jubilee National Hospital, Clydebank; Birmingham Clinical Trials Unit – University of Birmingham; University Hospitals Birmingham; Department of Inflammation and Ageing, University of Birmingham, and University Hospitals Birmingham NHS Foundation Trust; Manchester University NHS Foundation Trust, Wythenshawe Hospital, and School of Health and Society, University of Salford; Sheffield Centre for Health and Related Research, University of Sheffield; Warwick Clinical Trials Unit, Warwick Medical School, University of Warwick and Heartlands Good Hope Solihull Hospitals, University Hospitals Birmingham NHS Foundation Trust; FMedSci University of Birmingham, and University Hospitals Birmingham, Edgbaston Birmingham

## Abstract

Thoracotomy surgery is associated with high levels of acute pain and can lead to chronic post-surgical pain. Two methods of regional analgesia are commonly used in patients undergoing thoracotomy surgery, thoracic epidural blockade (TEB) and paravertebral blockade (PVB). The economic evaluation aimed to compare PVB to TEB, in terms of both costs and outcomes, and assess the cost-effectiveness of the different analgesic techniques, considering both impacts on chronic pain and health-related quality of life.

TOPIC-2 was a prospective, multi-centre, randomised, open-label, parallel-group, superiority trial of 770 adult (≥18 years old) thoracotomy patients, recruited from secondary care. The main economic analysis aimed to assess cost-effectiveness based on incremental cost per quality-adjusted life year (QALY) gained at 12 months post-randomisation, with a secondary analysis of cost per case of chronic post-thoracotomy pain (CPTP) avoided at 6 months.

The cost-utility analysis demonstrated that TEB resulted in slightly more QALYs at 12 months and was slightly more costly than PVB; however, the differences in costs and outcomes were not statistically significant at the 95% level. The results the sensitivity analyses showed that there was considerable uncertainty around the difference in costs and outcomes between the treatment groups.

The results of the analysis support the findings of other studies which suggest that there is no difference in the incidence of CPTP between patients receiving PVB compared with TEB and provides additional insights on health-related quality of life and costs. The study provides important information around chronic post-operative pain for policy makers, clinicians and patients.

## 1. Introduction

Thoracotomy surgery is associated with high levels of acute pain and can lead to chronic post-surgical pain, which can last for months and even years [36]. Each year, approximately 7,200 thoracotomies are conducted in the UK [46]. Chronic post-thoracotomy pain (CPTP) is defined as pain that recurs or persists for at least three months following surgery, and CPTP has been found to affect up to half of all patients undergoing this procedure [8; 46]. CPTP has been found to be associated with more frequent primary care visits and lower levels of productivity, alongside impacts on quality of life and other aspects of health [9; 37; 45]. Therefore, CPTP has important impacts for patients, healthcare services and society more broadly.

Two methods of regional analgesia are commonly used in patients undergoing thoracotomy surgery, thoracic epidural blockade (TEB) and paravertebral blockade (PVB). Compared to TEB, which blocks nerves in the chest bilaterally at the spinal cord level, some suggest that by unilaterally blocking nerve transmission more laterally, at the paravertebral space, PVB could be uniquely effective in preventing long-term pain [12; 33; 34]^-^. Both methods of pain control have been shown to be effective in managing pain in the acute postoperative period, where good analgesia is believed to be important to facilitate early mobilisation and physiotherapy, in turn reducing the risk of post-operative complications [14; 17; 46]. There is limited current evidence however, to support the most effective choice of anaesthetic technique in preventing CPTP, and therefore current UK practice varies greatly [3]. TEB has been historically regarded as the ‘gold standard’ for analgesia during thoracotomy, however, increasingly PVB is being employed [15; 38; 40]. In our pilot feasibility trial conducted in preparation for the presented study, the incidence of CPTP six-months postoperatively appeared lower with PVB compared than with TEB but a definitive trial was required to confirm this finding reliably [46].

The TOPIC2 trial tested the hypothesis that in patients undergoing thoracotomy, the use of PVB for postoperative pain relief reduces the 6-month incidence of CPTP compared with TEB [39]. This study presents the economic evaluation which was performed alongside the clinical trial. The economic evaluation aimed to compare PVB to TEB, in terms of both costs and outcomes, and assess the cost-effectiveness of the different analgesic techniques, considering both impacts on chronic pain and health-related quality of life.

## 2. Methods

### 2.1 Trial design and participants

Full details of the trial are published elsewhere (add reference). In brief, TOPIC-2 was a prospective, multi-centre, randomised, open-label, parallel-group, superiority trial of 770 adult (≥18 years old) thoracotomy patients, recruited from secondary care. Participants were recruited from 15 thoracic centres in the UK between January 2019 and September 2023.

Participants were eligible if they were: aged 18 years and above, undergoing elective open thoracotomy, able to provide documented informed consent to participate, and willing to complete trial questionnaires up until 12 months post-randomisation. Participants were excluded if they had: any contraindication to TEB or PVB (e.g., known allergy to local anaesthetics, infection near the proposed puncture site, coagulation disorders, thoracic spine disorders), rib/chest wall resection or planned pleurectomy, previous thoracotomy on the same side, or median sternotomy within 90 days.

Patients were randomised in a 1:1 ratio to either TEB (standard treatment) or PVB (interventional treatment). Randomisation was provided by a secure online randomisation system at the Birmingham Clinical Trials Unit (BCTU) using a minimisation algorithm within the online randomisation system to ensure balance in the treatment allocation over the following variables: gender, age < 65 years or ≥ 65 years, centre, and thoracotomy for lung cancer resection or for other indication.

All participants provided written informed consent and were followed up one month after the surgery. Follow-up continued at six and twelve months after randomisation. The trial was approved by the Southeast Scotland Research Ethics Committee, (REC 18/SS/0131) and registered with ClinicalTrials.gov, NCT03677856. The trial protocol has been published [39].

### 2.2 Economic evaluation overview

The economic evaluation used the data collected within the trial, and estimates of cost-effectiveness aimed to include the main outcome within the trial, which was CPTP at six months post-randomisation defined as a score of ≥40 when patients described their ‘worst chest pain over the last week’ on a 100mm visual analogue scale (VAS). The main economic analysis aimed to assess cost-effectiveness based on incremental cost per quality adjusted life year (QALY) gained at 12 months post-randomisation, with a secondary analysis of cost per case of CPTP avoided at 6 months. The primary economic analysis was conducted from a National Health Service (NHS) and Personal Social Services (PSS) perspective, based on cost per additional quality-adjusted life-year (QALY) gained. The methods used for this within-trial analysis were guided by UK National Institute for Health and Care Excellence (NICE) recommendations [19], and data analysis was carried out using Stata (version 18.0). Findings were reported in accordance with the Consolidated Health Economic Evaluation Reporting Standards (CHEERS) guidelines [22]. The time-horizon for the economic analysis was 12 months; therefore, costs and outcomes were not discounted. All costs are shown in British pounds (2021/2).

### 2.3 Resource use and costs

In the base-case analysis, resource costs were measured from a UK NHS perspective. Resource-use data was collected using case report forms (CRFs), completed by the trial staff until primary hospital discharge, and by the patients at 3, 6, and 12 months. A range of Information on resource use was collected such as the number of primary care service visits, NHS walk in centres and pharmacy consultations. Other healthcare resource use (outpatient, accident and emergency services) as well as additional treatments were also reported. Resource use data included: (1) the randomised analgesic intervention (PVB or TEB); (2) thoracotomy or the surgical procedure; (3) management in the acute phase, including post-operative management of the local anaesthetic block, analgesia, ward care, critical care, and, any additional theatre visits; (4) primary care and community-based services, including contacts with primary and community healthcare personnel (general practitioner, practice nurse, physiotherapist, psychologist, counsellor, pain specialist, district nurse, acupuncturist, osteopath, chiropractor, and other healthcare contacts); (5) inpatient hospital admissions (readmissions), including theatre visits; (6) emergency department visits; (7) medications; (8) equipment; and (9) other miscellaneous expenses, such as productivity costs incurred by the patients, as well as any private costs. Private health costs incurred by patients, and productivity costs were considered in the sensitivity analysis.

Unit costs were obtained from different sources, including but not confined to Personal Social Services Research Unit (PSSRU) Costs [3], NHS Reference Costs [1], British National Formulary (BNF) [2], NHS tariff book [4] and online sources, and Annual Survey of Hours and Earnings (ASHE) [31]. The unit costs are shown in Supplementary Tables. The micro-costing approach was employed to estimate the total costs for different cost categories by multiplying the resource item by the unit cost and summing all the items. All resource use was valued in monetary terms using appropriate UK unit costs estimated at the time of analysis. All costs were valued in UK currency (£) for 2024.

### 2.4 Health outcomes

The primary health outcome for the TOPIC2 trial was the presence of CPTP at 6 months post-randomisation. For this outcome, participants were asked to indicate their ‘worst chest pain over the last week’ on a 100-mm visual analogue scale (VAS, 0-100). Following published evidence, the presence of CPTP was defined as a VAS score which was greater or equal to 40mm, which has been reported as indicating at least a moderate level of pain [29]. The outcome measure for the main economic analysis, a cost-utility analysis (CUA) was QALYs. In line with NICE recommendations, health-related quality of life (HRQoL) was measured using the EuroQol 5-dimensions 5-level questionnaire (EQ-5D-5L) instrument [16]. The EQ-5D instrument is a preference-based measure consisting of five dimensions: mobility, self-care, usual activities, pain/discomfort, and anxiety/depression. The EQ-5D-5L instrument is widely used and is validated for patients with chronic pain (e.g.[30]). Each patient’s health status descriptions obtained from the EQ-5D-5L were translated into a single, preference-based (utility) index using a UK-specific value set [32]. Following NICE guidance, utility values were calculated by mapping the 5L descriptive system data onto the 3L value set[18]. We used the mapping function developed by van Hout et al. for the analysis, to allow consistency with NICE recommendations [43]. This mapping function uses data obtained from a survey of the UK population to derive a utility-based value [32]. Following the trapezium rule, the generated score was used to calculate Quality-Adjusted Life Years (QALYs) gained at different time points from baseline to 12 months after randomisation [27].

### 2.5 Base case analysis

For the base case analysis both a cost-utility analysis and a cost-effectiveness analysis were planned, with incremental cost-effectiveness ratios (ICER) estimated where appropriate. The analysis was conducted on an intention-to-treat (ITT) basis, following a pre-agreed health economics analysis plan (HEAP). Cost and outcome data were summarised and reported as mean values and standard deviations (SD), and resource use data were reported as medians and interquartile ranges (IQR). Given that cost data are likely to be positively skewed, 95% confidence intervals (CI) around differences in costs and outcomes from 1000 resamples were measured using the bias-corrected (BC) bootstrap method [6].

Incremental cost-effectiveness ratios (ICERs) were calculated based on the cost per QALY gained. The findings were interpreted using NICE’s willingness to pay (WTP) thresholds where an intervention is usually considered as cost-effective if the generated ICER is less than £20,000 – £30,000 per QALY gained [6].

### 2.6 Sensitivity analyses

A number of deterministic sensitivity analyses were carried out to assess the impact of variation in the estimated values and assumptions on the base case results. Plausible ranges were specified using information from the clinical trial and the literature. To allow consideration of a broader (societal) perspective costs incurred by participants and their families were included in the sensitivity analysis. This included private health costs incurred by patients, and productivity costs were also considered in the sensitivity analysis to assess the robustness of the findings. In addition, multiple imputation by chained equation [24; 44] and predictive mean matching were employed to replace missing cost data and EQ-5D-5L scores, assuming that values were missing at random (MAR), by generating fifty imputed datasets and combining them using Rubin’s rule [11; 35; 41]. To account for uncertainty due to sampling, a probabilistic sensitivity analyses comprising a non-parametric bootstrapping approach was applied to the patient level data to derive 5000 paired estimates of mean differences in costs and health outcomes [20]. The paired estimates were presented as scatterplots for the cost utility analyses on a cost effectiveness plane to facilitate the interpretation [10].

## 3. Results

Overall, 1327 participants were screened for eligibility between January 2019 and September 2023, and 770 patients were randomised to the PVB arm (n=386) or the TEB arm (n=384). Of those randomised, 33 did not proceed to thoracotomy, and withdrawal and follow up rates were similar across both groups.

### 3.1 Resource use and costs

Table 1 shows the aggregated total NHS costs for the participants. Details of the individual cost categories are in Supplementary Tables 1-3 and 9. The patients randomised to the TEB group had slightly higher health resource costs than those in the PVB group. The mean (SD) total NHS cost per patient, 12 months after randomisation, was £16,216.94 (£8,697.59) in the TEB group, and £16,137.27 (£10,154.76) in the PVB group, but these differences were not statistically significant. Thoracotomy costs contributed the most (around 52-53%) to the total NHS costs in both groups, followed by acute phase costs (around 41-42%).

**Table 1:** Aggregated costs, 12 months from randomisation, for all cost categories – base case, NHS perspective (UK£, 2021/22)

| Cost categories | PVB |  |  | TEB |  |  | Mean adjusted bootstrapped difference in costs (95% CI) |  |
| --- | --- | --- | --- | --- | --- | --- | --- | --- |
|  | N | Mean | SD | N | Mean | SD | Coefficient | P-value |
| Intervention | 386 | 530.16 | 524.58 | 384 | 518.01 | 328.96 | -12.15 | - |
| Operation | 386 | 8413.53 | 1979.37 | 384 | 8589.49 | 1490.20 | 175.96 (-47.08, 445.03) | 0.165 |
| Acute phase day 0 | 386 | 34.47 | 309.12 | 384 | 10.39 | 107.82 | -24.08 (-60.23, 4.92) | 0.141 |
| Acute phase day 1-3 | 386 | 2932.72 | 1414.34 | 384 | 3047.48 | 1284.91 | 114.75 (-82.22, 319.41) | 0.251 |
| Acute phase day 4-discharge | 386 | 3684.80 | 8180.82 | 384 | 3780.07 | 7479.60 | 95.27 (-985.74, 1208.81) | 0.866 |
| Primary care, physiotherapy, psychologist, counsellor visits etc. | 386 | 36.97 | 244.56 | 384 | 28.60 | 100.05 | -8.37 (-38.67, 12.38) | 0.525 |
| Inpatient readmissions at 3, 6, and 12 months post randomisation | 386 | 465.69 | 3495.10 | 384 | 208.40 | 1635.27 | -257.29 (-721.80, 60.30) | 0.180 |
| Emergency department visits | 386 | 16.93 | 77.14 | 384 | 13.87 | 68.56 | -3.06 (-13.38, 6.54) | 0.557 |
| Medications | 386 | 21.84 | 41.15 | 384 | 20.07 | 39.41 | -1.76 (-7.80, 4.06) | 0.558 |
| Equipment | 386 | 0.17 | 2.46 | 384 | 0.56 | 7.15 | 0.40 (-0.14, 1.39) | 0.287 |
| **Total NHS costs** | **386** | **16137.27** | **10154.76** | **384** | **16216.94** | **8697.592** | **79.66 (-1326.89, 1428.07)** | **0.909** |

### 3.2 Outcomes

The primary outcome of the trial was CPTP at six months. Overall, fifty-nine (22%) of the 272 participants in the PVB group and 47 (16%) of 292 participants in the TEB group reached this endpoint (adjusted RR 1·32 [95% CI 0·93 to 1·86]; adjusted RD 0·05 [95% CI –0·01 to 0·11]; p=0·12; Table 2). The analysis found that there was no significant difference in the primary outcome (‘worst pain in chest in the last week’, VAS ≥ 40) at six months. As no significant difference in this outcome was identified, the planned cost-effectiveness analysis, in terms of cost per case of CPTP avoided, was not undertaken.

**Table 2:**
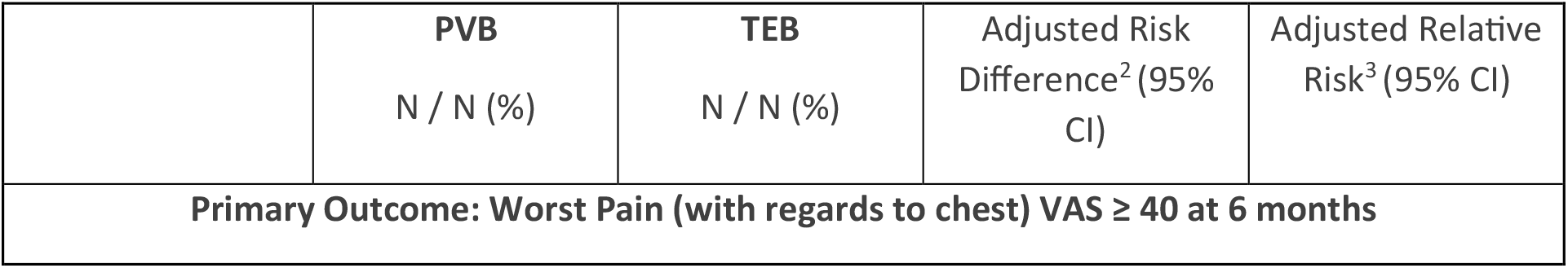
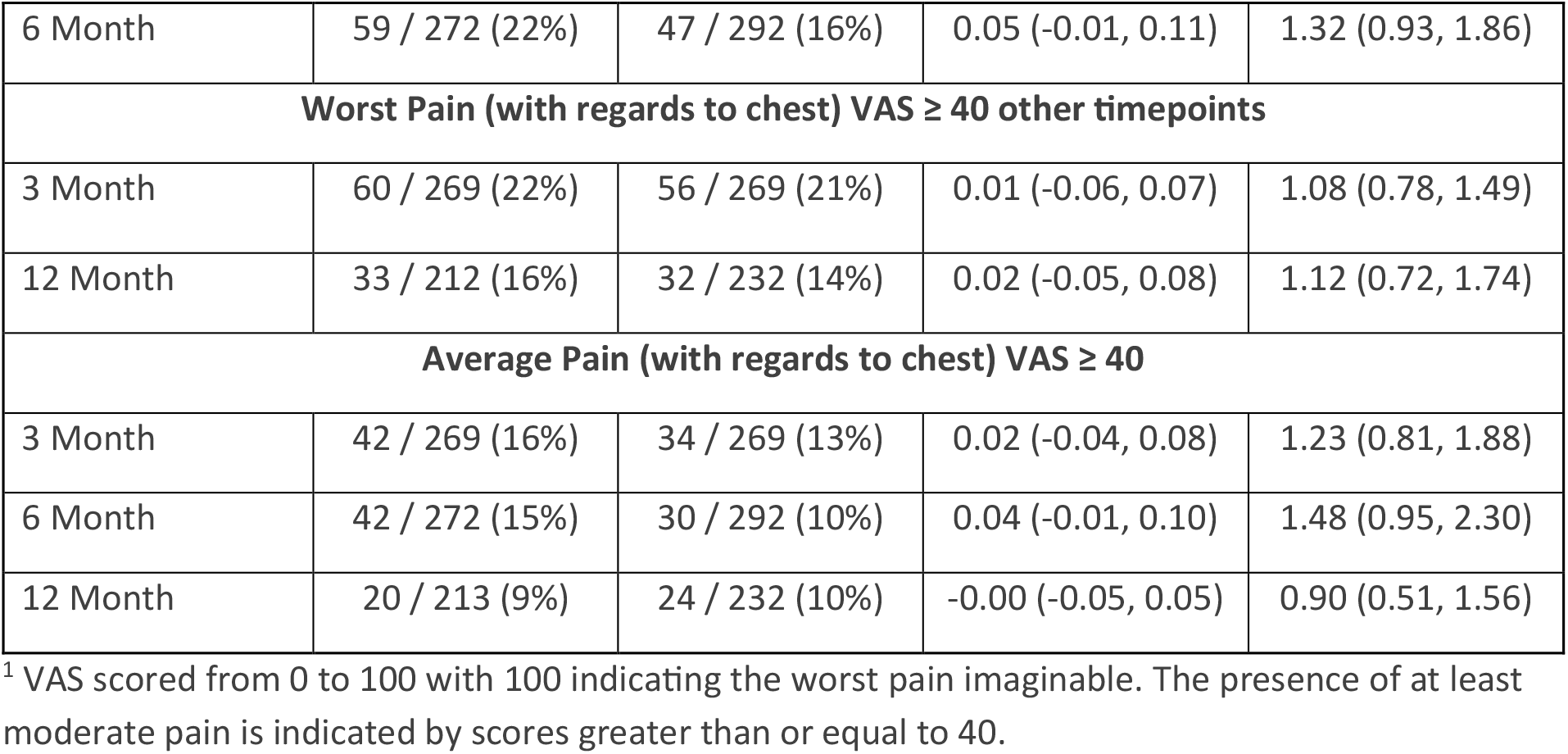
VAS^1^ Chronic Pain Responses.

The utility values for the two groups are shown in Table 3. The utility values were slightly higher for the PVB group at baseline compared to the TEB group, but up to 12 months, the participants randomised to the TEB group reported slightly higher utility values than those randomised to the PVB group. However, the differences in the utility values were not statistically significant apart from at 12 months.

**Table 3:** Utility values at different follow-up points.

| Utility values | PVB |  |  | TEB |  |  | Mean adjusted bootstrapped difference in utility values (95% CI) |  |
| --- | --- | --- | --- | --- | --- | --- | --- | --- |
|  | N | Mean | SD | N | Mean | SD | Coefficient | P-value |
| Baseline | 377 | 0.80 | 0.20 | 374 | 0.79 | 0.19 | -0.01 (-0.04, 0.02) | 0.595 |
| Discharge | 255 | 0.59 | 0.23 | 275 | 0.61 | 0.22 | 0.02 (-0.02, 0.06) | 0.297 |
| 3 months | 269 | 0.68 | 0.24 | 272 | 0.69 | 0.23 | 0.01 (-0.03, 0.05) | 0.581 |
| 6 months | 265 | 0.67 | 0.26 | 277 | 0.70 | 0.23 | 0.03 (-0.01, 0.07) | 0.118 |
| 12 months | 216 | 0.67 | 0.28 | 231 | 0.72 | 0.22 | 0.06 (0.01, 0.10) | 0.016 |

The mean QALY gain for the two groups at various time points is shown in Table 4. It is evident that generally statistically significant differences were not observed between the two groups, apart from between 6 and 12 months. At the 12 month point, the overall QALY gain for the TEB group was higher than for the PVB group, however, this was not found to be statistically significant.

**Table 4:** QALYs at different follow-up points.

| QALYs | PVB |  |  | TEB |  |  | Mean adjusted bootstrapped difference in QALYs (95% CI) |  |
| --- | --- | --- | --- | --- | --- | --- | --- | --- |
|  | N | Mean | SD | N | Mean | SD | Coefficient | P-value |
| From baseline to discharge | 255 | 0.02 | 0.01 | 271 | 0.03 | 0.12 | 0.01 (0.00, 0.04) | 0.131 |
| From baseline to 3 months | 268 | 0.19 | 0.05 | 268 | 0.19 | 0.04 | 0.00 (-0.01, 0.01) | 0.644 |
| From 3 month to 6 month | 241 | 0.17 | 0.06 | 245 | 0.18 | 0.05 | 0.01 (0.00, 0.02) | 0.149 |
| From 6 month to 12 month | 202 | 0.34 | 0.12 | 219 | 0.36 | 0.10 | 0.02 (0.00, 0.05) | 0.032 |
| **Overall (from baseline to 12 months)** | **150** | **0.73** | **0.21** | **166** | **0.77** | **0.24** | **0.04 (-0.01, 0.09)** | **0.153** |

### 3.3 Cost-utility analysis

Table 5 shows details of the cost-utility (CUA) analyses. In the base case analysis, the CUA showed that costs and QALY gains for PVB and TEB were very similar, but that TEB was associated with slightly higher costs and QALYs when compared to PVB. The average cost per participant in the PVB arm was £16,137.27 compared with £16,216.94 for the TEB arm, and the QALY gain for the PVB arm was 0.73, compared with 0.77 for the TEB arm (see Supplementary Table 4 for further information). The cost-utility analysis demonstrated that TEB resulted in slightly more QALYs at 12 months and was slightly more costly than PVB; however, the differences in costs and outcomes were not statistically significant at the 95% level. This resulted in an incremental cost-effectiveness ratio (ICER) of £1,991.50 per QALY gained, however, this needs to be considered in light of the sensitivity analyses below.

**Table 5:** CUA results – base case, NHS perspective at 12 months.

|  | Mean Cost (SD) | Incremental Cost (£) | Mean Outcome in QALYs (SD) | Incremental QALYs | ICER (Cost per QALY) |
| --- | --- | --- | --- | --- | --- |
| ICERs using NHS costs |  |  |  |  |  |
| PVB | 16,137.27<br>(10154.76) |  | 0.73 (0.21) |  |  |
| TEB | 16,216.94<br>(8697.59) | 79.66 | 0.77 (0.24) | 0.04 | 1,991.50 |

**Table 6:** Aggregated costs, 12 months from randomisation, for all cost categories – sensitivity analysis, societal perspective.

| Cost categories | PVB |  |  | TEB |  |  | Mean adjusted bootstrapped difference in costs (95% CI) |  |
| --- | --- | --- | --- | --- | --- | --- | --- | --- |
|  | N | Mean | SD | N | Mean | SD | Coefficient | P-value |
| Total NHS costs | 386 | 16137.27 | 10154.76 | 384 | 16216.94 | 8697.592 | 79.66 (-1326.89, 1428.07) | 0.909 |
| Productivity costs | 386 | 322.54 | 1758.68 | 384 | 300.24 | 2275.44 | -22.29 (-302.48, 276.34) | 0.874 |
| Private costs | 386 | 5.12 | 56.55 | 384 | 4.37 | 25.28 | -0.75 (-8.41, 4.45) | 0.815 |
| **Total societal costs** | **386** | **16464.94** | **10288.54** | **384** | **16521.55** | **8990.364** | **56.61 (-1374.91, 1392.52)** | **0.937** |

### 3.4 Sensitivity analyses

The results of 5000 bootstrap replications plotted on the cost-effectiveness plane for the primary analysis (Figure 1) shows that there is uncertainty around the difference in costs between the treatment arms, as replicates are distributed almost equally across the north-west quadrant (less effective and greater cost) and south-west quadrant (less effective and lower cost). In addition, although the majority of the bootstrap replicates were located in the north-west and south-west quadrant, suggesting that TEB is more costly and more effective than PVB at 12 months, considerable uncertainty was apparent, as many replicates were found in the north-east quadrant (more effective and more costly) and in the south-west quadrant (less effective and less costly).

**Figure 1:**
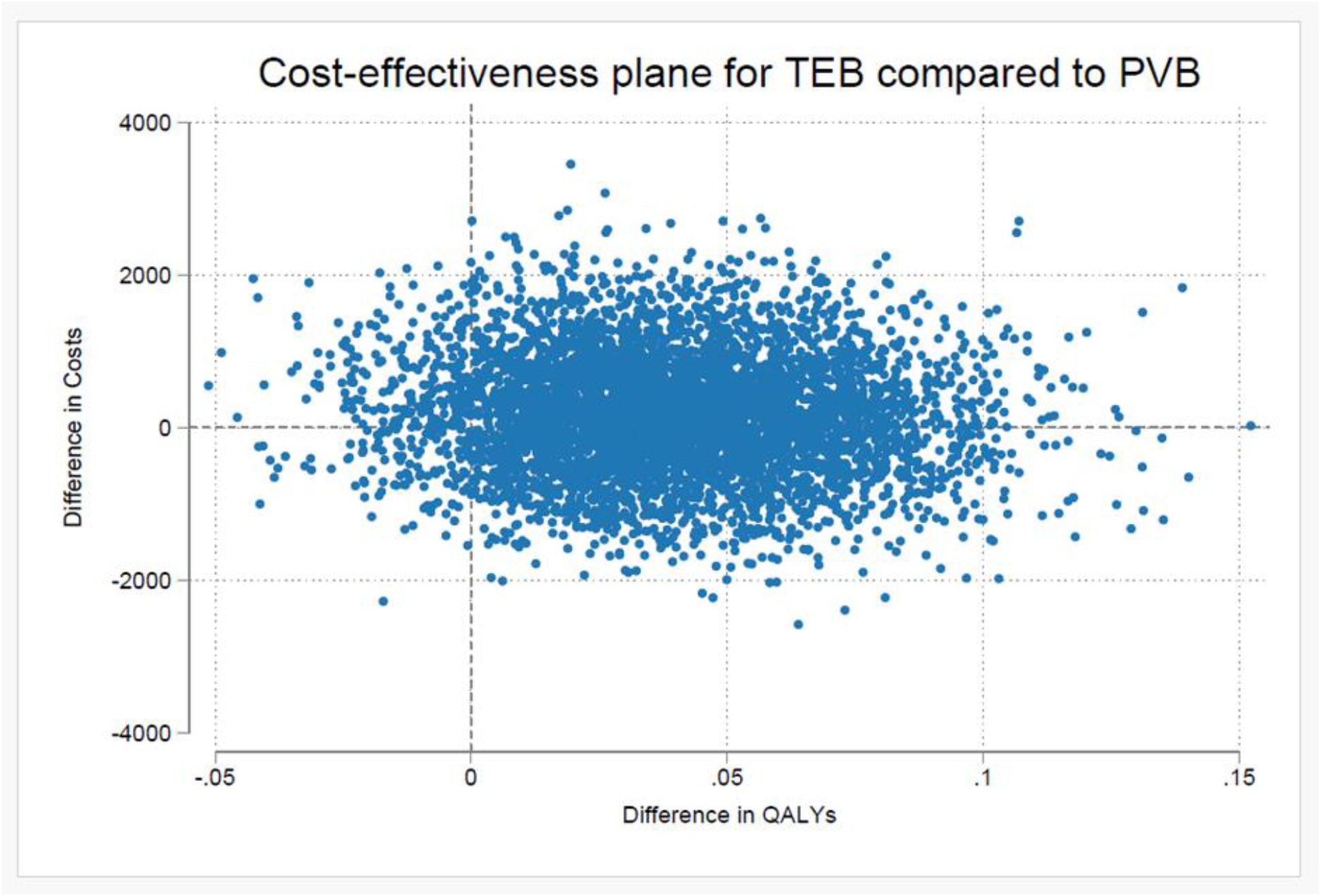
Cost-effectiveness plane for TEB compared to PVB (12 months)

The results of the deterministic sensitivity analyses which explored the impact of varying the assumptions for different scenarios are shown below. Including broader costs for the participants groups did not affect the study results, as broader costs for both study groups were similar. Supplementary Table 5 shows the aggregated total societal costs for the participants. Details of the individual cost categories are in Supplementary Table 5. The mean (SD) total societal cost per patient, 12 months after randomisation, was £16,521.55 (£8,990.36) in the TEB group, and £16,464.94 (£10,288.54) in the PVB group. TEB was associated with higher costs (£56.61) and QALYs (0.06) when compared to PVB, and with an ICER of £943.50 per QALY gained, however, these differences were not statistically significant. The results of employing multiple imputation methods to handle missing data are shown in Supplementary Tables 6-7.

## 4. Discussion

This trial-based economic evaluation analysed the cost-effectiveness of PVB compared with TEB in the UK for people undergoing thoracotomy. The analyses showed that TEB was associated with a lower number of participants reporting experiencing worst chest pain at the 6-month follow-up point in the TEB group, compared with the PVB group, but that this difference was not statistically significant. The cost-utility analysis demonstrated that TEB produced an additional 0.04 QALYs when compared to PVB, and was associated with higher costs, but these differences were not statistically significant. The probabilistic sensitivity analysis demonstrated that there was considerable uncertainty around the estimates of differences in costs and outcomes between the study arms. The deterministic sensitivity analysis showed that the inclusion of productivity and private costs did not change the results of the evaluation.

This is the first health economic analysis of PVB compared with TEB in thoracotomy patients in the UK, adopting both a healthcare and societal perspective. The study adds to the limited cost-effectiveness literature relating to anaesthesia [25; 42]. In particular, there has been very little focus on chronic post-operative pain, with previous studies in this perioperative medicine focusing on the reduction of perioperative infection[13], prevention of delirium[21], and postoperative care [26]. The study adds to the growing literature on the cost-effectiveness of interventions concerned with chronic pain (e.g. [5; 7]). The study also provides a focus on post-operative chronic pain, which is a less examined area within cost-effectiveness studies [23; 28].

This study has several strengths. Firstly, the economic evaluation was conducted alongside a rigorously conducted randomised controlled trial across 15 centres in all 4 nations of the UK. The analysis addresses some of the limitations associated with previous analyses in similar areas, comprehensively incorporating evidence on both clinical outcomes and health-related quality of life, as well as the adoption of a 12-month follow-up period. In addition, a societal perspective was explored as a secondary analysis which gives additional depth to the evaluation. Moreover, there is a lack of economic evidence relating to different analgesic techniques and associated impacts on chronic pain, and hence this study makes an important contribution where evidence is generally lacking.

There were some limitations associated with this analysis. In particular, there was a lack of detail associated with the information from participants in relation to the use of painkillers and other medications, which meant that a range of assumptions needed to be adopted. These assumptions were informed by detailed discussions with clinicians, and extensive sensitivity analyses were undertaken. In addition, the study’s conclusions are based on the NHS healthcare system and may not be applicable to other healthcare systems. For example, in settings where substantially different approaches to anaesthesia and staffing exist, different results might be encountered. Similarly, the analyses are based on currently available methods, and they may not be applicable to future surgical techniques. In addition, there were some limitations associated with the trial itself, as there was some loss to follow up, and blinding of participants and clinicians to the study intervention was not possible.

This study has important implications for research and policy both in the UK and internationally. Firstly, the results of the analysis support the findings of other studies which suggest that there is no difference in the incidence of CPTP between patients receiving PVB compared with TEB. This study also suggests that there is no evidence of a significant difference in terms of health-related quality of life and costs. The study provides information for policy makers, clinicians and patients and can add to the information provided to patents around chronic post-operative pain. The study additionally highlights the need for more evidence around different approaches to anaesthesia and the impacts on health-related quality of life and post-operative pain. In view of the TOPIC-2 trial suggesting no significant differences in terms of the clinical nor cost effectiveness between TEB and PVB, both techniques are likely to continue to be used, allowing appropriate deployment when the clinical situation demands, and facilitating individualised analgesic plans to be made for each patient.

## Data Availability

The datasets generated during the current study will be made available by the Chief Investigator upon reasonable request and in accordance with the Birmingham Clinical Trials Unit's research collaboration and data transfer guidelines.

## Conflict of Interest Statement

Funding for this study was provided by National Institute for Health and Care Research (NIHR) 16/111/111

The authors have no conflicts of interest to declare.

## Notes

### Competing Interest Statement

The authors have declared no competing interest.

### Clinical Trial

NCT03677856

### Funding Statement

This project was funded by the National Institute for Health and Care Research (NIHR) Health
Technology Assessment (HTA) programme (NIHR 16/111/111)

### Author Declarations

Ethics committee of South-East Scotland Research Ethics Committee (REC 18/SS/0131) gave ethical approval for this work.

