## Supplementary Materials for "The cost-effectiveness of thoracic epidural versus paravertebral blockade in reducing chronic post-thoracotomy pain – a trial-based economic evaluation"

***Supplementary Table 1: Unit costs of anaesthetics and analgesics***

| Anaesthetics and analgesics | Available form/dose | Size | Cost of vials/bottles/ box | Cost per unit |
|---|---|---|---|---|
| **Pre-medication to recovery** | | | | |
| IV Paracetamol | 1g/100 ml | 10 bottles/vials | £ 12.45 | £ 1.25 |
| IV NSAIDs – Diclofenac | 75mg/3ml | 10 ampoules | £ 9.91 | £ 0.99 |
| IV NSAIDS – Ketorolac | 30mg/1ml | 5 ampoules | £ 5.36 | £ 1.07 |
| Oral Codeine | 30mg | 28 tablets | £ 0.98 | £ 0.04 |
| IV Tramadol | 100mg/2ml | 5 ampoules | £ 4.00 | £ 0.80 |
| IV Remifentanil | 1mg | 5 vials | £ 25.50 | £ 5.10 |
| IV Alfentanil | 1mg/2ml | 10 ampoules | £ 9.00 | £ 0.90 |
| IV Fentanyl | 2.5mg/50ml | 1 vial | £ 9.80 | £ 9.80 |
| Topical Fentanyl | 12µg/1hr | 5 patches | £ 5.64 | £ 1.13 |
| IV Morphine | 50mg/50ml | 1 vial | £ 5.78 | £ 5.78 |
| Oral Morphine | 10mg | 56 tablets | £ 5.31 | £ 0.09 |
| IV Diamorphine | 5mg | 5 ampoules | £ 12.80 | £ 2.56 |
| IV Oxycodone | 10mg/1ml | 5 ampoules | £ 6.80 | £ 1.36 |
| Oral Oxycodone | 5mg | 20 tablets | £ 1.95 | £ 0.10 |
| Oral Pregabalin | 25mg | 56 tablets | £ 3.99 | £ 0.07 |
| Oral Gabapentin | 600mg | 100 tablets | £ 6.30 | £ 0.06 |
| IV Ketamine | 200mg/20ml | 1 vial | £ 5.06 | £ 5.06 |
| **Analgesic intervention** | | | | |
| Bupivacaine Infusion | 250mg/250ml (0.1%) | 5 bags | £ 60.87 | £ 12.17 |
| Bupivacaine Injection | 25mg/10ml (0.25%) | 5 ampoules | £ 9.99 | £ 2.00 |
| Levobupivacaine Infusion | 125mg/100ml (0.125%) | 5 bags | £ 51.85 | £ 10.37 |

| | | | | |
|---|---|---|---|---|
| Levobupivacaine Injection | 25mg/10ml (0.25%) | 5 ampoules | £ 10.58 | £ 2.12 |
| Ropivacaine Infusion | 400mg/200ml (0.2%) | 5 bags | £ 78.50 | £ 15.70 |
| Ropivacaine Injection | 20mg/10ml (0.2%) | 5 ampoules | £ 12.65 | £ 2.53 |
| Fentanyl Infusion | 2.5mg/50ml (0.005%) | 1 vial | £ 9.80 | £ 9.80 |
| Fentanyl Injection | 100µg/2ml (0.005%) | 10 ampoules | £ 11.50 | £ 1.15 |
| Morphine (preservative-free) Infusion | 100mg/100ml (0.1%) | 1 bag | £ 23.13 | £ 23.13 |
| Morphine (preservative-free) Injection | 10mg/10ml (0.1%) | 10 ampoules | £ 15.00 | £ 1.50 |
| Diamorphine Infusion/Injection | 5mg | 5 ampoules | £ 12.80 | £ 2.56 |
| **Post-operative analgesia (acute phase)** | | | | |
| Co-codamol | 30mg/500mg | 100 tablets | £2.94 | £0.03 |
| Oral Codeine | 30mg | 28 tablets | £0.98 | £0.04 |
| IV Tramadol | 100mg/2ml | 5 ampoules | £4.00 | £0.80 |
| Oral Tramadol | 100mg | 60 tablets | £6.94 | £0.12 |
| IV Fentanyl | 2.5mg/50ml | 1 vial | £9.80 | £9.80 |
| Topical Fentanyl | 25µg/1hr | 5 patches | £8.07 | £1.61 |
| IV Morphine | 50mg/50ml | 1 vial | £5.78 | £5.78 |
| Oral Morphine | 20mg | 56 tablets | £10.61 | £0.19 |
| Oral Oxycodone | 5mg | 20 tablets | £1.95 | £0.10 |
| Oral Pregabalin | 75mg | 56 tablets | £4.79 | £0.09 |
| Oral Gabapentin | 600mg | 100 capsules | £6.30 | £0.06 |
| IV Ketamine | 200mg/20ml | 1 vial | £5.06 | £5.06 |
| **Post-operative analgesia (from discharge till 12 months)** | | | | |
| Paracetamol | 1g | 100 tablets | £9.20 | £0.09 |

| | | | | |
|---|---|---|---|---|
| Codeine | 30mg | 28 tablets | £0.98 | £0.04 |
| Gabapentin | 300mg | 100 capsules | £2.32 | £0.02 |
| Ibuprofen | 400mg | 60 tablets | £4.90 | £0.08 |
| Diclofenac sodium | 1% | 100g | £4.17 | £0.04 |
| Zopiclone | 7.5mg | 28 tablets | £1.13 | £0.04 |
| Citalopram | 10mg | 28 tablets | £0.64 | £0.02 |
| Lidocaine | 5% | 30 plasters | £61.54 | £2.05 |

***Supplementary Table 2: Unit costs of healthcare personnel, inpatient admissions, theatre and emergency department visits, and surgical intervention and procedures***

| **Resource use item** | **Unit cost** | **Description** | **Source** |
|---|---|---|---|
| **Healthcare personnel** | | | |
| Anaesthetist | £143.00 | Hospital-based doctors (consultant: surgical), cost per working hour (including qualifications) | PSSRU 2022 Table 11.3.2 |
| Consultant surgeon | £143.00 | Hospital-based doctors (consultant: surgical), cost per working hour (including qualifications) | PSSRU 2022 Table 11.3.2 |
| Trainee surgeon | £73.00 | Hospital-based doctors (registrar), cost per working hour (including qualifications) | PSSRU 2022 Table 11.3.2 |
| GP | £41.00 | GP, per surgery consultation lasting 9.22 minutes (with qualification costs) | PSSRU 2022 Table 9.4.2 |
| Practice nurse | £52.00 | GP practice nurse (band 5), cost per working hour (including qualifications) | PSSRU 2022 Table 9.3.1 |
| Physiotherapist | £55.00 | Community-based scientific and professional staff (band 6), cost per working hour | PSSRU 2022 Tables 8.1 and 8.2.1 |
| Psychologist | £66.00 | Community-based scientific and professional staff (band 7), cost per working hour | PSSRU 2022 Tables 8.1 and 8.2.1 |
| Counsellor | £55.00 | Community-based scientific and professional staff (band 6), cost per working hour | PSSRU 2022 Tables 8.1 and 8.2.1 |
| Pain specialist (community) | £77.10 | Community Health Services - Other therapist, adult, one to one - A01A1 | NHS Reference Costs 2021-22 |
| District nurse | £63.00 | Qualified nurse (band 6), cost per working hour (including qualifications) | PSSRU 2022 Table 9.2.1 |

| | | | |
|---|---|---|---|
| Acute nurse | £59.00 | Hospital-based nurses (band 6), cost per working hour (including qualifications) | PSSRU 2022 Table 11.2.2 |
| Acupuncturist | £77.10 | Community Health Services - Other therapist, adult, one to one - A01A1 | NHS Reference Costs 2021-22 |
| Osteopath | £77.10 | Community Health Services - Other therapist, adult, one to one - A01A1 | NHS Reference Costs 2021-22 |
| Chiropractor | £77.10 | Community Health Services - Other therapist, adult, one to one - A01A1 | NHS Reference Costs 2021-22 |
| Other | £77.10 | Community Health Services - Other therapist, adult, one to one - A01A1 | NHS Reference Costs 2021-22 |
| **Inpatient admissions, and theatre and emergency department visits** | | | |
| General ward | £633.94 | Weighted average of Unspecified Pain, Unspecified Chest Pain, Abdominal Pain & Observation or Counselling - (WH08A-WH08B, EB12A-EB12C, FD05A-FD05B, WH16A-WH16B) - Elective, Non-elective long stay, Non-elective short stay & Daycase | NHS Reference Costs 2021-22 |
| Acute ward | £415.88 | Weighted average of Unspecified Pain, Unspecified Chest Pain, Abdominal Pain & Observation or Counselling - (WH08A-WH08B, EB12A-EB12C, FD05A-FD05B, WH16A-WH16B) - Non-elective short stay | NHS Reference Costs 2021-22 |
| HDU - acute phase | £1,502.66 | Thoracic Surgical Adult Patients Predominate, 0 Organs Supported, XC07Z - Critical Care | NHS Reference Costs 2021-22 |
| HDU - 3m/6m/12m | £1,709.19 | Non-specific, General Adult Patients Predominate, 0 Organs Supported, XC07Z - Critical Care | NHS Reference Costs 2021-22 |
| ITU - acute phase | £1,812.08 | Weighted average of Thoracic Surgical Adult Patients Predominate, 1-2 Organs Supported, XC05Z-XC06Z - Critical Care | NHS Reference Costs 2021-22 |
| ITU - 3m/6m/12m | £2,293.15 | Weighted average of Non-specific, General Adult Patients Predominate, 1-6+ Organs Supported, XC01Z-XC06Z - Critical Care | NHS Reference Costs 2021-22 |
| Theatre visits | £349.98 | Weighted average of Thoracic Surgery Service, Consultant Led, Non-Admitted & Multipprofessional Face-to-Face | NHS Reference Costs 2021-22 |

| | | | |
|---|---|---|---|
| | | Attendance, WF01B & WFO2B - Outpatient Care | |
| Emergency department visits | £242.02 | Weighted average of No Investigation with No Treatment & Any Category Investigation with Any Category Treatment, VB01Z-VB09Z, VB11Z - Emergency Care | NHS Reference Costs 2021-22 |
| **Surgical intervention and procedures** | | | |
| Operation | £8,493.06 | Weighted average of Major, Complex & Very Complex Thoracic Procedures, 19 years and over - (DZ63A-DZ63C, DZ02H-DZ02K, DZ62A-DZ62C) - Total HRGs | NHS Reference Costs 2021-22 |
| Bronchoscopy | £1,479.83 | Weighted average of Therapeutic & Diagnostic Bronchoscopy - (DZ68Z, DZ69A) - Total HRGs | NHS Reference Costs 2021-22 |
| Redo thoracotomy | £3,041.18 | Weighted average of Minor, Intermediate & Major Thoracic Procedures, 19 years and over - (DZ71Z, DZ64A-DZ64C, DZ63A-DZ63C) - Total HRGs | NHS Reference Costs 2021-22 |

***Supplementary Table 3: Unit costs of intraoperative monitoring devices, investigations and procedures, equipment, and time off work***

| **Resource use item** | **Unit cost** | **Description** | **Source** |
|---|---|---|---|
| **Intraoperative monitoring** | | | |
| Central line/central venous catheter | £9.11 | Prosys Vesica Community Catheterisation Pack, | https://www.nhsbsa.nhs.uk/sites/default/files/2022-06/Drug%20Tariff%20Part%20IX%20July%202022.pdf, pg 411) |
| Urinary catheter | £2.30 | Foley Catheter, For Short/medium Term Use in Adults, | (https://www.nhsbsa.nhs.uk/sites/default/files/2022-06/Drug%20Tariff%20Part%20IX%20July%202022.pdf, pg 423) |
| Arterial line | £6.99 | 20g 45mm BD Arterial Cannula Floswitch, | (https://ukmedi.co.uk/products/20g-45mm-bd-arterial-cannula-floswitch-bd-682245-ukmedi-co-uk) |
| **Investigations and procedures** | | | |

| | | | |
|---|---|---|---|
| X-ray | £28.38 | Weighted average of Plain Film, PF IMAGDA and IMAGOP - IMAG | NHS Reference Costs 2021-22 |
| CT scan | £123.42 | Weighted average of Computerised Tomography Scan of One Area, 19 years and above (RD20A, RD21A, RD22Z) - Total HRGs | NHS Reference Costs 2021-22 |
| MRI | £207.16 | Weighted average of Magnetic Resonance Imaging Scan of One Area, 19 years and above (RD01A, RD02A, RD03Z) - Total HRGs | NHS Reference Costs 2021-22 |
| Chest drain | £15.19 | Chest Drain | (https://www.nhsbsa.nhs.uk/sites/default/files/2022-06/Drug%20Tariff%20Part%20IX%20July%202022.pdf, pg 525) |
| **Equipment** | | | |
| Bath seat | £9.00 | Bath seat | https://livingmadeeasy.org.uk/category/health-and-personal-care/bathing-and-toileting/bathshower-boards-seats-and-stools/product/ailsa-bath-seat |
| Electronic mattress support | £125.93 | Mattress tilter | https://www.medicalsupplies.co.uk/MGS.html?gad_source=1&gclid=Cj0KCQjw99e4BhDiARIsAISE7P-biYuLyZY2KK3mqFirSdT7VPaDQNX3hidjrYpxZC60ZdE0KZT2O1YaAhN_EALw_wcB |
| Rollator | £35.00 | Steel three wheel rollator walker | https://livingmadeeasy.org.uk/category/access-and-mobility/walking-aids/rollators-and-triwalkers/product/steel-three-wheel-rollator-walker |
| TENS machine | £20.00 | TENS machine | https://www.nhs.uk/conditions/transcutaneous-electrical-nerve-stimulation-tens/ |
| Walking stick | £5.00 | Wooden walking stick | https://livingmadeeasy.org.uk/category/access-and-mobility/walking- |

| | | | |
|---|---|---|---|
| | | | aids/walking-sticks-and-canes/product/chestnut-wooden-walking-stick |
| Hand rails for bathroom and stairs | £6.49 | Straight household grab bar | https://www.screwfix.com/p/essentials-straight-household-abs-grab-bar-white-300mm/646FH?tc=EH6&gad_source=1&gclid=Cj0KCQjw99e4BhDiARIsAISE7P-dWy_QbSTnYmhQir-4OmunDb7LUjF6U9oQjy9BFnl2mr5KkwnGzxkaAtU_EALw_wcB&gclsrc=aw.ds |
| Seat with back rest | £53.00 | Back suport | https://livingmadeeasy.org.uk/product/backfriend-back-support |
| **Productivity** | | | |
| Male - full time employee | £857.90 | Average weekly pay | ASHE 2023 |
| Female - full time employee | £728.00 | Average weekly pay | ASHE 2023 |
| Male - part time employee | £270.40 | Average weekly pay | ASHE 2023 |
| Female - part time employee | £284.70 | Average weekly pay | ASHE 2023 |

***Supplementary Table 4: Disaggregated costs, 12 months from randomisation, for all cost categories – base case***

| **Cost categories** | **PVB** | | | **TEB** | | | **Mean adjusted bootstrapped difference in costs (95% CI)** | |
|---|---|---|---|---|---|---|---|---|
| | **N** | **Mean** | **SD** | **N** | **Mean** | **SD** | **Coefficient** | **P-value** |
| Anaesthetist costs | 303 | 598.26 | 210.50 | 302 | 604.49 | 219.23 | 6.23 (-28.52, 39.41) | 0.712 |
| Total pre-medication to recovery costs | 386 | 40.455 | 459.085 | 384 | 15.501 | 140.351 | -24.954 (-84.541, 11.290) | 0.307 |
| Analgesic intervention costs | 386 | 11.06 | 8.19 | 384 | 16.62 | 9.63 | 5.56 | - |
| Total intraoperative monitoring costs | 386 | 9.04 | 4.72 | 384 | 10.49 | 5.02 | 1.45 (0.75, 2.14) | 0.000 |

| | | | | | | | | |
|---|---|---|---|---|---|---|---|---|
| **Total intervention costs** | **386** | **530.16** | **524.58** | **384** | **518.01** | **328.96** | **-12.15** | **-** |
| Thoracotomy costs | 386 | 8053.01 | 1884.93 | 384 | 8249.77 | 1418.57 | 196.76 (15.36, 465.27) | 0.103 |
| Surgeon costs | 378 | 368.16 | 212.07 | 371 | 351.62 | 199.82 | -16.53 (-45.29, 13.48) | 0.279 |
| **Total operation costs** | **386** | **8413.53** | **1979.37** | **384** | **8589.49** | **1490.20** | **175.96 (-47.08, 445.03)** | **0.165** |
| Management of local anaesthetic costs | 386 | 1.40 | 8.97 | 384 | 2.45 | 17.03 | 1.06 (-0.66, 3.31) | 0.282 |
| Post-operative analgesic costs | 386 | 0.65 | 2.28 | 384 | 0.22 | 1.39 | -0.42 (-0.69, -0.16) | 0.002 |
| Theatre costs | 386 | 32.42 | 308.80 | 384 | 7.71 | 106.66 | -24.71 (-60.98, 3.68) | 0.130 |
| **Total day 0 costs** | 386 | 34.47 | 309.12 | 384 | 10.39 | 107.82 | -24.08 (-60.23, 4.92) | 0.141 |
| Management of local anaesthetic costs | 386 | 3.05 | 13.71 | 384 | 5.52 | 19.51 | 2.47 (0.41, 5.11) | 0.037 |
| Post-operative analgesic costs | 386 | 0.57 | 1.89 | 384 | 0.35 | 1.46 | -0.22 (-0.45, 0.02) | 0.077 |
| Theatre costs | 386 | 48.66 | 346.12 | 384 | 16.75 | 219.85 | -31.91 (-72.82, 6.40) | 0.119 |
| Ward care costs | 386 | 2880.44 | 1329.60 | 384 | 3024.85 | 1241.11 | 144.41 (-45.10, 331.09) | 0.135 |
| **Total day 1-3 costs** | 386 | 2932.72 | 1414.34 | 384 | 3047.48 | 1284.91 | 114.75 (-82.22, 319.41) | 0.251 |
| Management of local anaesthetic costs | 386 | 3.14 | 12.21 | 384 | 2.50 | 10.97 | -0.64 (-2.19, 1.08) | 0.441 |
| Post-operative analgesic costs | 386 | 5.64 | 17.30 | 384 | 6.02 | 15.30 | 0.38 (-1.77, 2.81) | 0.745 |
| Theatre costs | 386 | 27.75 | 271.41 | 384 | 27.40 | 296.98 | -0.35 (-43.58, 40.20) | 0.987 |
| Ward care costs | 386 | 3648.27 | 8133.26 | 384 | 3744.15 | 7410.45 | 95.88 (-982.22, 1207.15) | 0.865 |

| | | | | | | | | |
|---|---|---|---|---|---|---|---|---|
| **Total day 4-discharge costs** | 386 | 3684.80 | 8180.82 | 384 | 3780.07 | 7479.60 | 95.27 (-985.74, 1208.81) | 0.866 |
| GP costs | 386 | 21.14 | 61.22 | 384 | 15.27 | 42.30 | -5.87 (-13.54, 1.50) | 0.118 |
| Practice nurse costs | 386 | 2.29 | 29.78 | 384 | 4.74 | 47.52 | 2.45 (-2.22, 9.70) | 0.394 |
| Physiotherapist costs | 386 | 0.43 | 4.84 | 384 | 0.00 | 0.00 | -0.43 (-1.07, -0.14) | 0.058 |
| Psychologist costs | 386 | 0.00 | 0.00 | 384 | 0.00 | 0.00 | - | - |
| Counsellor costs | 386 | 0.00 | 0.00 | 384 | 0.00 | 0.00 | - | - |
| Pain specialist (community) costs | 386 | 0.00 | 0.00 | 384 | 2.61 | 47.37 | 2.61 (0.19, 7.71) | 0.231 |
| District nurse costs | 386 | 8.32 | 153.99 | 384 | 1.97 | 38.58 | -6.36 (-23.67, 4.76) | 0.417 |
| Acupuncturist costs | 386 | 0.00 | 0.00 | 384 | 0.00 | 0.00 | - | - |
| Osteopath costs | 386 | 0.00 | 0.00 | 384 | 0.00 | 0.00 | - | - |
| Chiropractor costs | 386 | 0.00 | 0.00 | 384 | 0.00 | 0.00 | - | - |
| Other costs | 386 | 4.79 | 36.55 | 384 | 4.02 | 25.82 | -0.78 (-5.72, 3.39) | 0.739 |
| **Total healthcare personnel costs** | **386** | **36.97** | **244.56** | **384** | **28.60** | **100.05** | **-8.37 (-38.67, 12.38)** | **0.525** |
| General ward costs | 386 | 448.36 | 3453.23 | 384 | 183.25 | 1229.61 | -265.11 (-708.63, 21.83) | 0.143 |
| Acute/short stay costs | 386 | 3.23 | 36.57 | 384 | 5.42 | 87.45 | 2.18 (-4.52, 15.01) | 0.660 |
| HDU costs | 386 | 0.00 | 0.00 | 384 | 0.00 | 0.00 | - | - |
| ITU costs | 386 | 5.94 | 116.72 | 384 | 17.92 | 351.07 | 11.97 (-17.68, 54.31) | 0.548 |
| Theatre costs | 386 | 8.16 | 58.59 | 384 | 1.82 | 35.72 | -6.34 (-13.26, 0.28) | 0.064 |
| **Total inpatient admission costs** | **386** | **465.69** | **3495.10** | **384** | **208.40** | **1635.27** | **-257.29 (-721.80, 60.30)** | **0.180** |
| **Emergency department visits costs** | **386** | **16.93** | **77.14** | **384** | **13.87** | **68.56** | **-3.06 (-13.38, 6.54)** | **0.557** |

| | | | | | | | | |
|---|---|---|---|---|---|---|---|---|
| Conventional painkillers costs | 386 | 13.72 | 26.08 | 384 | 12.11 | 23.69 | -1.61 (-5.15, 1.88) | 0.379 |
| Opioids costs | 386 | 4.40 | 10.35 | 384 | 4.54 | 11.38 | 0.14 (-1.50, 1.70) | 0.864 |
| Neuropathic painkillers costs | 386 | 0.66 | 2.32 | 384 | 0.64 | 2.34 | -0.02 (-0.39, 0.28) | 0.888 |
| Anti-inflammatory drugs costs | 386 | 1.27 | 5.35 | 384 | 0.62 | 4.59 | -0.64 (-1.31, 0.13) | 0.073 |
| Gels/creams costs | 386 | 0.72 | 7.14 | 384 | 0.56 | 7.22 | -0.16 (-1.20, 0.85) | 0.756 |
| Sleeping pills costs | 386 | 0.04 | 0.37 | 384 | 0.05 | 0.50 | 0.01 (-0.05, 0.08) | 0.707 |
| Anti-depressants costs | 386 | 0.10 | 0.70 | 384 | 0.01 | 0.13 | -0.09 (-0.17, -0.03) | 0.014 |
| Patches costs | 386 | 0.93 | 12.17 | 384 | 1.55 | 15.00 | 0.62 (-1.32, 2.53) | 0.538 |
| **Total medication costs** | **386** | **21.84** | **41.15** | **384** | **20.07** | **39.41** | **-1.76 (-7.80, 4.06)** | **0.558** |
| **Total equipment costs** | **386** | **0.17** | **2.46** | **384** | **0.56** | **7.15** | **0.40 (-0.14, 1.39)** | **0.287** |
| **Total NHS costs** | **386** | **16137.27** | **10154.76** | **384** | **16216.94** | **8697.592** | **79.66 (-1326.89, 1428.07)** | **0.909** |

*Supplementary Table 5: Disaggregated costs, 12 months from randomisation, for all cost categories – sensitivity analysis*

| **Cost categories** | **PVB** | | | **TEB** | | | **Mean adjusted bootstrapped difference in costs (95% CI)** | |
|---|---|---|---|---|---|---|---|---|
| | **N** | **Mean** | **SD** | **N** | **Mean** | **SD** | **Coefficient** | **P-value** |
| Anaesthetist costs | 303 | 598.26 | 210.50 | 302 | 604.49 | 219.23 | 6.23 (-28.52, 39.41) | 0.712 |
| Total pre-medication to recovery costs | 386 | 40.455 | 459.085 | 384 | 15.501 | 140.351 | -24.954 (-84.541, 11.290) | 0.307 |
| Analgesic intervention costs | 386 | 11.06 | 8.19 | 384 | 16.62 | 9.63 | 5.56 | - |

| | | | | | | | | |
|---|---|---|---|---|---|---|---|---|
| Total intraoperative monitoring costs | 386 | 9.04 | 4.72 | 384 | 10.49 | 5.02 | 1.45 (0.75, 2.14) | 0.000 |
| **Total intervention costs** | **386** | **530.16** | **524.58** | **384** | **518.01** | **328.96** | **-12.15** | **-** |
| Thoracotomy costs | 386 | 8053.01 | 1884.93 | 384 | 8249.77 | 1418.57 | 196.76 (15.36, 465.27) | 0.103 |
| Surgeon costs | 378 | 368.16 | 212.07 | 371 | 351.62 | 199.82 | -16.53 (-45.29, 13.48) | 0.279 |
| **Total operation costs** | **386** | **8413.53** | **1979.37** | **384** | **8589.49** | **1490.20** | **175.96 (-47.08, 445.03)** | **0.165** |
| Management of local anaesthetic costs | 386 | 1.40 | 8.97 | 384 | 2.45 | 17.03 | 1.06 (-0.66, 3.31) | 0.282 |
| Post-operative analgesic costs | 386 | 0.65 | 2.28 | 384 | 0.22 | 1.39 | -0.42 (-0.69, -0.16) | 0.002 |
| Theatre costs | 386 | 32.42 | 308.80 | 384 | 7.71 | 106.66 | -24.71 (-60.98, 3.68) | 0.130 |
| **Total day 0 costs** | 386 | 34.47 | 309.12 | 384 | 10.39 | 107.82 | -24.08 (-60.23, 4.92) | 0.141 |
| Management of local anaesthetic costs | 386 | 3.05 | 13.71 | 384 | 5.52 | 19.51 | 2.47 (0.41, 5.11) | 0.037 |
| Post-operative analgesic costs | 386 | 0.57 | 1.89 | 384 | 0.35 | 1.46 | -0.22 (-0.45, 0.02) | 0.077 |
| Theatre costs | 386 | 48.66 | 346.12 | 384 | 16.75 | 219.85 | -31.91 (-72.82, 6.40) | 0.119 |
| Ward care costs | 386 | 2880.44 | 1329.60 | 384 | 3024.85 | 1241.11 | 144.41 (-45.10, 331.09) | 0.135 |
| **Total day 1-3 costs** | 386 | 2932.72 | 1414.34 | 384 | 3047.48 | 1284.91 | 114.75 (-82.22, 319.41) | 0.251 |
| Management of local anaesthetic costs | 386 | 3.14 | 12.21 | 384 | 2.50 | 10.97 | -0.64 (-2.19, 1.08) | 0.441 |
| Post-operative analgesic costs | 386 | 5.64 | 17.30 | 384 | 6.02 | 15.30 | 0.38 (-1.77, 2.81) | 0.745 |
| Theatre costs | 386 | 27.75 | 271.41 | 384 | 27.40 | 296.98 | -0.35 (-43.58, 40.20) | 0.987 |

| | | | | | | | | |
|---|---|---|---|---|---|---|---|---|
| Ward care costs | 386 | 3648.27 | 8133.26 | 384 | 3744.15 | 7410.45 | 95.88 (-982.22, 1207.15) | 0.865 |
| **Total day 4-discharge costs** | 386 | 3684.80 | 8180.82 | 384 | 3780.07 | 7479.60 | 95.27 (-985.74, 1208.81) | 0.866 |
| GP costs | 386 | 21.14 | 61.22 | 384 | 15.27 | 42.30 | -5.87 (-13.54, 1.50) | 0.118 |
| Practice nurse costs | 386 | 2.29 | 29.78 | 384 | 4.74 | 47.52 | 2.45 (-2.22, 9.70) | 0.394 |
| Physiotherapist costs | 386 | 0.43 | 4.84 | 384 | 0.00 | 0.00 | -0.43 (-1.07, -0.14) | 0.058 |
| Psychologist costs | 386 | 0.00 | 0.00 | 384 | 0.00 | 0.00 | - | - |
| Counsellor costs | 386 | 0.00 | 0.00 | 384 | 0.00 | 0.00 | - | - |
| Pain specialist (community) costs | 386 | 0.00 | 0.00 | 384 | 2.61 | 47.37 | 2.61 (0.19, 7.71) | 0.231 |
| District nurse costs | 386 | 8.32 | 153.99 | 384 | 1.97 | 38.58 | -6.36 (-23.67, 4.76) | 0.417 |
| Acupuncturist costs | 386 | 0.00 | 0.00 | 384 | 0.00 | 0.00 | - | - |
| Osteopath costs | 386 | 0.00 | 0.00 | 384 | 0.00 | 0.00 | - | - |
| Chiropractor costs | 386 | 0.00 | 0.00 | 384 | 0.00 | 0.00 | - | - |
| Other costs | 386 | 4.79 | 36.55 | 384 | 4.02 | 25.82 | -0.78 (-5.72, 3.39) | 0.739 |
| **Total healthcare personnel costs** | **386** | **36.97** | **244.56** | **384** | **28.60** | **100.05** | **-8.37 (-38.67, 12.38)** | **0.525** |
| General ward costs | 386 | 448.36 | 3453.23 | 384 | 183.25 | 1229.61 | -265.11 (-708.63, 21.83) | 0.143 |
| Acute/short stay costs | 386 | 3.23 | 36.57 | 384 | 5.42 | 87.45 | 2.18 (-4.52, 15.01) | 0.660 |
| HDU costs | 386 | 0.00 | 0.00 | 384 | 0.00 | 0.00 | - | - |
| ITU costs | 386 | 5.94 | 116.72 | 384 | 17.92 | 351.07 | 11.97 (-17.68, 54.31) | 0.548 |
| Theatre costs | 386 | 8.16 | 58.59 | 384 | 1.82 | 35.72 | -6.34 (-13.26, 0.28) | 0.064 |
| **Total inpatient admission costs** | **386** | **465.69** | **3495.10** | **384** | **208.40** | **1635.27** | **-257.29 (-721.80, 60.30)** | **0.180** |

| | | | | | | | | |
|---|---|---|---|---|---|---|---|---|
| **Emergency department visits costs** | **386** | **16.93** | **77.14** | **384** | **13.87** | **68.56** | **-3.06 (-13.38, 6.54)** | **0.557** |
| Conventional painkillers costs | 386 | 13.72 | 26.08 | 384 | 12.11 | 23.69 | -1.61 (-5.15, 1.88) | 0.379 |
| Opioids costs | 386 | 4.40 | 10.35 | 384 | 4.54 | 11.38 | 0.14 (-1.50, 1.70) | 0.864 |
| Neuropathic painkillers costs | 386 | 0.66 | 2.32 | 384 | 0.64 | 2.34 | -0.02 (-0.39, 0.28) | 0.888 |
| Anti-inflammatory drugs costs | 386 | 1.27 | 5.35 | 384 | 0.62 | 4.59 | -0.64 (-1.31, 0.13) | 0.073 |
| Gels/creams costs | 386 | 0.72 | 7.14 | 384 | 0.56 | 7.22 | -0.16 (-1.20, 0.85) | 0.756 |
| Sleeping pills costs | 386 | 0.04 | 0.37 | 384 | 0.05 | 0.50 | 0.01 (-0.05, 0.08) | 0.707 |
| Anti-depressants costs | 386 | 0.10 | 0.70 | 384 | 0.01 | 0.13 | -0.09 (-0.17, -0.03) | 0.014 |
| Patches costs | 386 | 0.93 | 12.17 | 384 | 1.55 | 15.00 | 0.62 (-1.32, 2.53) | 0.538 |
| **Total medication costs** | **386** | **21.84** | **41.15** | **384** | **20.07** | **39.41** | **-1.76 (-7.80, 4.06)** | **0.558** |
| **Total equipment costs** | **386** | **0.17** | **2.46** | **384** | **0.56** | **7.15** | **0.40 (-0.14, 1.39)** | **0.287** |
| **Total NHS costs** | **386** | **16137.27** | **10154.76** | **384** | **16216.94** | **8697.592** | **79.66 (-1326.89, 1428.07)** | **0.909** |
| **Total productivity costs** | **386** | **322.54** | **1758.68** | **384** | **300.24** | **2275.44** | **-22.29 (-302.48, 276.34)** | **0.874** |
| Healthcare costs | 386 | 0.19 | 3.66 | 384 | 1.67 | 20.93 | 1.48 (-0.06, 4.26) | 0.160 |
| Admissions costs | 386 | 0.00 | 0.00 | 384 | 0.00 | 0.00 | - | - |
| Medicines costs | 386 | 1.48 | 6.67 | 384 | 1.79 | 10.26 | 0.31 (-0.83, 1.68) | 0.625 |
| Equipment costs | 386 | 3.46 | 56.11 | 384 | 0.92 | 7.32 | -2.54 (-10.21, 0.89) | 0.385 |
| **Total private costs** | **386** | **5.12** | **56.55** | **384** | **4.37** | **25.28** | **-0.75 (-8.41, 4.45)** | **0.815** |

| | | | | | | | | |
|---|---|---|---|---|---|---|---|---|
| **Total societal costs** | **386** | **16464.94** | **10288.54** | **384** | **16521.55** | **8990.364** | **56.61 (-1374.91, 1392.52)** | **0.937** |

**Supplementary Table 6: Sensitivity analysis (using multiple imputations)**

| **Total costs** | **PVB** | | | **TEB** | | | **Mean adjusted bootstrapped difference in costs (95% CI)** | |
|---|---|---|---|---|---|---|---|---|
| | **N** | **Mean** | **SD** | **N** | **Mean** | **SD** | **Coefficient** | **P-value** |
| Intervention costs | 386 | 626.21 | 186.77 | 384 | 636.86 | 195.25 | 10.65 | - |
| Operation costs | 386 | 8414.427 | 1979.771 | 384 | 8594.579 | 1490.349 | 180.15 (-44.11, 452.45) | 0.156 |
| Acute phase day 0 costs | 386 | 34.47 | 309.12 | 384 | 10.39 | 107.82 | -24.08 (-60.23, 4.92) | 0.141 |
| Acute phase day 1-3 costs | 386 | 2932.72 | 1414.34 | 384 | 3047.48 | 1284.91 | 114.75 (-82.22, 319.41) | 0.251 |
| Acute phase day 4-discharge costs | 386 | 3684.80 | 8180.82 | 384 | 3780.07 | 7479.60 | 95.27 (-985.74, 1208.81) | 0.866 |
| Primary care visits costs | 386 | 22.01 | 206.22 | 384 | 11.70 | 35.67 | -10.31 (-34.91, 3.99) | 0.327 |
| A&E costs | 386 | 18.83 | 76.94 | 384 | 14.99 | 68.47 | -3.85 (-14.28, 5.40) | 0.461 |
| Inpatient admissions costs | 386 | 465.69 | 3495.10 | 384 | 208.40 | 1635.27 | -257.29 (-721.80, 60.30) | 0.180 |
| Medication costs | 386 | 19.41 | 38.89 | 384 | 17.22 | 35.05 | -2.19 (-7.51, 3.27) | 0.420 |
| Equipment costs | 386 | 0.17 | 2.46 | 384 | 0.56 | 7.15 | 0.40 (-0.14, 1.39) | 0.287 |
| **Total NHS costs** | **386** | **16832.51** | **10136.16** | **384** | **16944.69** | **8691.03** | **112.18 (-1276.21, 1486.91)** | 0.873 |

**Supplementary Table 7: Sensitivity analysis (using multiple imputations) - ICER**

| | **Mean Cost (SD)** | **Incremental Cost** | **Mean Outcome (SD)** | **Incremental Outcome** | **ICER (Cost per outcome)** |
|---|---|---|---|---|---|
| **ICERs using NHS costs** | | | | | |
| PVB | 16832.51 (10136.16) | | 0.51 (0.36) | | |
| TEB | 16944.69 (8691.03) | 112.18 | 0.58 (0.35) | 0.07 | 1602.571429 |

**Supplementary Table 8: Assumptions**

| |
|---|
| Several assumptions were made when calculating the costs, which are listed below, for each category of resource use: |
| **Intervention** |
| • The intervention costs included: (1) anaesthetist costs; (2) analgesic (pre-medication to recovery) costs; (3) costs of the analgesic intervention itself (PVB or TEB); and (4) intraoperative monitoring costs.<br>• To calculate anaesthetist cost, the time duration from the time when the anaesthetist first met the patient in the theatre complex to the time the anaesthetist left theatre was considered.<br>• For calculating analgesic costs, the cost of the medication was obtained from BNF (NHS indicative price). Where the route of the medication was oral, the drug was assumed to be in tablet form, and the costs of tablets were considered. Where the route was patient-controlled analgesia (PCA), the medication was assumed to be intravenous, and the cost of intravenous drugs was considered.<br>• The costs of PVB included costs of single shot injections, loading of paravertebral catheters, analgesic infusion during and after surgery, and any top-ups required. The costs of TEB included costs of loading bolus, analgesic infusion during and after surgery, and any top-ups required. When calculating the cost of the analgesic intervention, the cost of injections was used, except for analgesic infusion, where the cost of intravenous form of drugs was used.<br>• The costs of intraoperative monitoring included the costs of arterial line, central venous catheter, and urinary catheter. Although an ultrasound machine is required with central venous line, the cost of the machine was not included. |
| **Operation** |
| • The operation costs included: (1) thoracotomy costs; and (2) consultant or trainee surgeon costs.<br>• Weighted average of major, complex, and very complex thoracic procedures was taken to determine thoracotomy costs.<br>• To calculate surgeon cost, the time duration from the time of knife to skin to the time the anaesthetist left theatre was considered. |
| **Acute phase** |

- The intervention costs included: (1) management of local anaesthesia costs; (2) post-operative analgesia costs; (3) return to theatre costs; and (4) post-operative ward care costs.
- For management of local anaesthesia, it was assumed that optimisation required either an anaesthetist or a nurse, and took about 20 minutes; while re-siting required both anaesthetist and nurse, and took about 40 minutes.
- Return to theatre costs included bronchoscopy, redo thoracotomy, and theatre visit costs. It was assumed that these procedures include the theatre and surgeon costs, and they were not costed separately. Also, for redo thoracotomy, it was assumed to be cheaper than the thoracotomy procedure itself, and weighted average of minor, intermediate, and major thoracic procedures was taken.

**Post-operative ward care and inpatient admissions**

- In the acute post-operative period, when calculating costs for the HDU and ITU, it was assumed that the patients were admitted where there were mostly adult patients with thoracic surgical conditions/procedures; while in the chronic phase, they were assumed to be admitted where there were mostly adult patients with non-specific conditions.
- The cost of ITU was assumed to be more than HDU.

**Analgesia and medications**

- The analgesics, form, concentration, and unit price have been tabulated elsewhere.
- When calculating the costs of the analgesics, the form, concentration and dose of drugs that were most frequently listed in the data was used. The costs were adjusted for each patient according to the concentration and dose.
- Where the dose, concentration or form of the medications was not mentioned, the following form, concentration, and doses were assumed:

| Analgesic | Form | Concentration | Dose |
|---|---|---|---|
| Gabapentin | Oral | 300mg | Twice a day |
| Pregabalin | Oral | 75mg | Twice a day |
| Ketamine | IV | 200mg | Two 200mg in 24h period |
| Codeine | Oral | 60mg | Four times a day |
| Tramadol | Oral | 100mg | Four times a day |
| Oxycodone | Oral | 40mg | Four times a day |
| Morphine | Oral | 40mg | Four times a day |
| Fentanyl | Topical | 25mcg/hour | One patch worn for 72h |

| |
|---|
| • Where only the class and duration of the days was mentioned, the following drugs, concentration, and doses were assumed |

| Category | Drug | Concentration | Dose |
|---|---|---|---|
| Conventional painkiller | Paracetamol | 1g | Four times a day |
| Opioids | Codeine | 30mg | Four times a day |
| Neuropathic painkillers | Gabapentin | 300mg | Twice a day |
| Anti-inflammatory drugs | Ibuprofen | 400mg | Thrice a day |
| Gels/creams | Diclofenac sodium | 1% | Four times a day |
| Sleeping pills | Zopiclone | 7.5mg | Once a day |
| Anti-depressants | Citalopram | 10mg | Once a day |
| Patches | Lidocaine | 5% | One patch worn for 12h |

| |
|---|
| **Equipment** |
| • For calculating cost of the equipment, the cheapest price available online was taken for the equipment listed in the data. |
| **Productivity** |
| • The average full-time and part-time salaries for both males and females was taken to get the average weekly pay, which was divided by 5 to get the daily wage, assuming 5 working days in a week. The daily wage was then multiplied by the number of days taken off work to calculate the productivity cost. |
| **Private** |
| • When calculating private costs, only the costs paid by the patients and their relatives/caregivers was considered, and not the costs covered by insurance or NHS. |
| **Miscellaneous** |
| • It was assumed that the costs of the investigations, like x-rays, and procedures, like chest drain, were included in the inpatient, theatre, and emergency department visits. |

**Supplementary Table 9: Non-Standard Abbreviations and Acronyms**

| | |
|---|---|
| ASHE | Annual Survey of Hours and Earnings |
| BC | Bias Corrected |
| BCTU | Birmingham Clinical Trials Unit |

| | |
|---|---|
| BNF | British National Formulary |
| BPI | Brief Pain Inventory |
| CEAC | Cost Effectiveness Acceptability Curve |
| CHEERS | Consolidated Health Economic Evaluation Reporting Standards |
| CI | Confidence Intervals |
| CPTP | Chronic Post-Thoracotomy Pain |
| CRF | Case Report Form |
| CUA | Cost Utility Analysis |
| EEPRU | Policy Research Unit in Economic Methods of Evaluation of Health and Care Interventions |
| EQ-5D-3L | EuroQol 5-Dimensions 3-Levels |
| EQ-5D-5L | EuroQol 5-Dimensions 5-Levels |
| HADS | Hospital Anxiety and Depression Scale |
| HEAP | Health Economics Analysis Plan |
| HRQoL | Health Related Quality of Life |
| ICER | Incremental Cost Effectiveness Ratio |
| ISRCTN | International Standard Randomised Controlled Trial Number |
| IQR | Interquartile Ranges |
| ITT | Intention To Treat |
| NHS | National Health Service |
| NICE | National Institute for Health and Care Excellence |
| NIHR | National Institute for Health and Care Research |
| PSS | Personal Social Services |
| PSSRU | Personal Social Services Research Unit |
| PVB | Paravertebral Blockade |
| QALY | Quality Adjusted Life Year |
| SD | Standard Deviations |

| | |
|---|---|
| SF-MPQ-2 | Short-Form McGill Pain Questionnaire-2 |
| TEB | Thoracic Epidural Blockade |
| TOPIC-2 | A Randomised Controlled Trial to Investigate the Effectiveness of Thoracic Epidural and Paravertebral Blockade In Reducing Chronic Post-Thoracotomy Pain: 2 |
| VAS | Visual Analogue Scale |
| WTP | Willingness To Pay |

**Supplementary Figure 1: Sensitivity analysis (using multiple imputations) – Cost-effectiveness Plane**

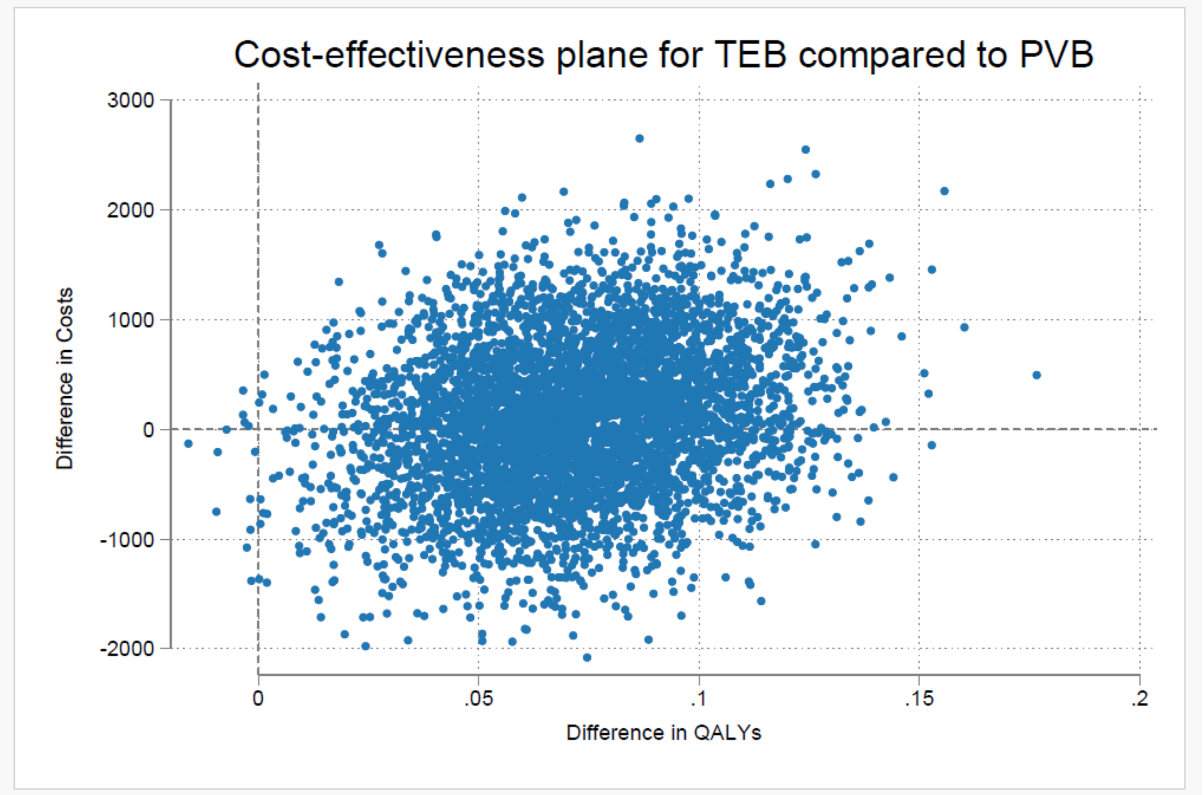
